# Predictors of COVID-19 Vaccine Hesitancy: Socio-demographics, Co-Morbidity and Past Experience of Racial Discrimination

**DOI:** 10.1101/2021.01.12.21249152

**Authors:** E. Savoia, R. Piltch-Loeb, B. Goldberg, C. Miller-Idriss, B. Hughes, AM. Montrond, JN. Kayyem, MA. Testa

## Abstract

**Importance:** Immunization programs are only successful when there are high rates of acceptance and coverage. While delivering billions of COVID 19 doses globally addressing vaccine hesitancy will be one of the most significant public health communication efforts ever undertaken.

**Objective:** The goal of this study is to explore predictors of COVID 19 vaccine hesitancy, including sociodemographic factors, comorbidity, risk perception, and experience of discrimination, in a sample of the U.S. population.

**Design:** We used a cross sectional online survey study design. The survey was implemented between Dec 13 and 23, 2020.

**Setting:** The survey was limited to respondents over 18 years of age residing in the USA.

**Participants:** Respondents were individuals belonging to priority groups for vaccine distribution.

**Main Outcome(s) and Measure(s):** Respondents were asked how likely they would be to take a COVID 19 vaccine if offered at no cost within two months. Vaccine hesitancy was measured using a scale ranging from 1 (low hesitancy) to 6 (high hesitancy).

**Results:** Responses were received from 2,650 respondents (response rate 84%) from all 50 states and Puerto Rico, American Samoa, and Guam. The majority were in the age category between 25 and 44 years (66%), male (53%), and working in the healthcare sector (61%). Most were White and non-Hispanic (66%) respondents followed by Black non-Hispanic (14%) and Hispanic (8%) respondents. Experience with racial discrimination was a predictor of vaccine hesitancy. Those reporting racial discrimination having 21% increased odds of being at a higher level of hesitancy compared to those who did not report such experience (OR=1.21, 95% C.I. 1.01-1.45).

**Conclusions and Relevance:** Communication and logistical aspects during the COVID 19 vaccination campaign need to be sensitive to individuals past-experience of discrimination by identifying appropriate channels of communication and sites for vaccine distribution to reach those who may have sentiments of mistrust in the vaccination campaign.

## Introduction

Within a year, the SARS-CoV-2 pandemic spread worldwide infecting millions of individuals and causing thousands of deaths. Under the federal Operation Warp Speed program, administered by the U.S. Department of Health and Human Services, $10 billion dollars were invested in six candidate vaccines.^1^ In November 2020, Pfizer-BioNTech^2^ and Moderna^3^ reported that their much-anticipated vaccines demonstrated over 90% effectiveness in protecting people from the disease. Both vaccines were developed and tested at record speed and given U.S. Food and Drug Administration (FDA) emergency use authorization (EUA) in December 2020. ^4,5^

Vaccine distribution of the Pfizer/BioNTech vaccine began on December 8 2020, starting from a 90 year-old grandmother in Great Britain. Two months prior to the approval of the vaccines, the Centers for Disease Control and Prevention (CDC) released its interim playbook for jurisdictional operations outlining a phased approach to COVID-19 vaccination starting from those considered most at risk due to their job, age and health status. ^6^

While in some ways the difficult scientific efforts in producing the vaccine have proven so successful, the delivery of the vaccine to the public is expected to face vast logistical, distribution and information systems challenges. Along with organizational aspects, hesitancy of individuals to take the vaccine is also a formidable challenge. Immunization programs are only successful when there are high rates of acceptance and coverage. Addressing vaccine hesitancy, while delivering billions of doses across the world will be one of the greatest public health risk communication efforts ever undertaken. As such, it is critical to understand the reasons why specific segments of the population are more hesitant than others to accept the vaccination, and address the reasons of such hesitancy to the extent possible when implementing distribution plans.^7^

Various opinion polls have identified high levels of vaccine skepticism and specific reasons for such skepticism. Kreps and colleagues^8^ found that vaccine efficacy and safety are important factors associated with public acceptance of a COVID-19 vaccine, as well as the process by which a vaccine is authorized and by whom it is endorsed. A survey of a representative sample of the U.S. population, conducted in May 2020, prior to the release of the Pfizer-BioNTech and Moderna vaccines, reported that 33% of the population was vaccine hesitant and that in particular Black respondents were less likely to accept a potential COVID-19 vaccine. ^9^ These results are fairly similar across polls, with some variation depending on the time the poll was conducted and the socio-demographics, co-morbidity characteristics of the sampled population. According to a recent longitudinal probability-based internet survey of the US population the likelihood of getting a COVID-19 vaccine declined from 74% in early April to 56% in early December 2020 ^10^. A more recent poll showed that 35% of Black respondents are hesitant about receiving the vaccine, 71% of which say they are worried about possible side effects, and 50% believe that they can become infected with COVID-19 from the vaccination.^11^ This low acceptance is consistent with historical disparities in influenza immunization behavior and perceptions in the U.S. population, with Black adults significantly less likely to receive the influenza vaccine than White adults.^12, 13^ This is particularly concerning considering that Black individuals shoulder a disproportionate burden of many chronic conditions, placing them at higher risk for complications from preventable diseases like influenza^14^, and now that vaccines are available, from COVID-19 as well. Research on the racial disparities in influenza immunization rates in the general population have identified several psychosocial and behavioral factors associated with vaccine uptake including: perceived risk, trust, vaccine attitudes, social norms, and experiences of racism.^15^ The goal of this study is to explore the predictors of COVID-19 vaccine hesitancy including socio-demographic factors, co-morbidity, risk perception and past-experience with discrimination, in particular among those identified as priority groups for the vaccination. The study is based on a rapid survey conducted around the time of the Pfizer-BioNTech and Moderna vaccines approval with the ultimate goal of informing public officials on how to enhance vaccine communication efforts during the vaccination campaign.

## Methods

### Study Design

We used a cross-sectional online survey study design. The survey was implemented via mobile phones by the use of the survey platform Pollfish, and it was limited to respondents over 18 years of age residing in the USA. Similar to third-party advertising companies, Pollfish pays mobile application developers to display and promote the surveys to their users. To incentivize participation, relatively small monetary reimbursements are provided to randomly selected users who complete the surveys. An initial survey instrument draft was implemented for cognitive testing with 20 individuals, and the survey was subsequently revised after feedback to include 36 questions. Questions and response choices were kept short using “yes/no” or Likert-type and rating scales to facilitate completion by the use of mobile phones. The survey was launched on December 13 and closed on December 23, 2020. A screening question was used to identify respondents belonging to one of 19 job categories that were identified as priority groups for vaccine distribution based on national guidance available at the time of the survey.^16^ The study protocol and survey instrument were approved by the Harvard T.H. Chan School of Public Health Institutional Review Board. A copy of the survey instrument can be found in Appendix.

### Dependent Variable

Multivariable ordinal regression was undertaken to model the underlying construct of vaccine hesitancy measured by the creation of a Likert-type scale. Respondents were asked how likely they would be to take a COVID-19 vaccine if offered to them at no cost within two months. Answer options were ordered as follows: very likely (1), somewhat likely (2), would consider it after two months (3), not sure (4), somewhat unlikely (5), very unlikely (6). Results were interpreted with a range of values from 1 (low hesitancy) to 6 (high hesitancy).

### Independent Variables

Independent predictor variables included socio-demographics such as age, gender, race, level of education and employment status. Other predictors included job type (working in the healthcare sector *versus* other priority groups for vaccination), having had a diagnosis of COVID-19 (with no symptoms, mild or severe symptoms), clinical risk of severe consequences from COVID-19, risk perception of contracting the disease or infecting others, and past experience with discrimination. Risk perception was measured by asking respondents to report their level of concern with contracting COVID-19 at work, outside their work environment, and infecting family members or friends. A factor analysis was performed to assess the structure of the risk perception questions, and as a result a scale was created with scores ranging from 0 to 6, with lower values indicating lower risk perception. Kaiser–Meyer–Olkin (KMO) measure of sampling adequacy was used to test for the suitability of the data for factor analysis and Cronbach’s alpha to assess the reliability of the scale. The respondents’ clinical risk for severe consequences from COVID-19 was measured by asking about the underlying health conditions most frequently associated with severe disease or death (diabetes, cardiovascular disease, obesity, pulmonary disease, immunocompromised status, rheumatological condition, or cancer), responses were converted into a dichotomous variable describing presence of at least one comorbidity *versus* absence of comorbidities. Finally, respondents were asked about past experience with unfair treatment they attributed to their race, religion, gender or sexual orientation using an adaptation of the discrimination scale developed by Sternthal, M.J. et al.^17^ This scale includes six questions on unfair treatment experienced in the work environment, at school, by a police officer and by financial institutions (i.e. bank loan). The adaptation consisted of adding a question about unfair treatment by a physician or nurse and by limiting the cause of the unfair treatment to race, religion, gender, and sexual orientation.

### Statistical Analyses

We first performed descriptive statistics for each variable. We then applied simple and multiple ordinal regression models to study the association between the independent variables and COVID-19 vaccine hesitancy (dependent variable). We tested for bivariate associations between each predictor [age, gender, race, education, employment status, job type, having had a diagnosis of COVID-19, clinical risk profile, risk perception and experience of discrimination] and the dependent variable, by means of ordinal and logistic regression using a p-value < 0.05 as cut-off for inclusion of the independent variables in the multiple regression model. We tested the parallel regression assumption by means of the *Brant* test for the ordinal logistic model which did not show statistical significance. The Stata Statistical Software 16 was used.

## Results

### Socio-demographic characteristics of the study population

Responses were received from 2,650 respondents (response rate 84%) from all 50 states and the territories of Puerto Rico, American Samoa and Guam. Descriptive statistics are given in Table 1. The five most represented states were California (13%), New York (10%), Texas (7%), Florida (6%) and Pennsylvania (4%). Sixty-six percent of respondents were age 25–44 years with median age 37 years, 53% were male, and 61% were working in the healthcare sector. The majority of respondents were white and non-Hispanic (66%) and others were Black non-Hispanic (14%) and Hispanic (8%). Respondents were highly educated with 31% having a graduate-level degree, and 86% were employed at the time of the survey.

**Table 1.** Socio-demographic characteristics of the study population

| Age | N (%) |
| --- | --- |
| 18-24 | 354 (13.4%) |
| 25-34 | 841 (31.7%) |
| 35-44 | 906 (34.2%) |
| 45-54 | 339 (12.8%) |
| 55-64 | 152 (5.7%) |
| 65-74 | 47 (1.8%) |
| ≥75 | 11 (0.4%) |
| Gender | N (%) |
| Male | 1417 (53.5%) |
| Female | 1213 (45.8%) |
| Other | 20 (0.7%) |
| Education | N (%) |
| Less than high school | 92 (3.2%) |
| High school/GED | 539 (20.3%) |
| Some college | 579 (21.9%) |
| Bachelor's degree | 615 (23.3%) |
| Post-graduate degree | 825 (31.3%) |
| Race |  |
| White, non-Hispanic | 1754 (66.2%) |
| Black, non-Hispanic | 379 (14.3%) |
| Hispanic | 206 (7.8%) |
| Asian, non-Hispanic | 130 (4.9%) |
| 2+ races | 122 (4.6%) |
| Prefer not to say | 40 (1.5%) |
| Other | 19 (0.7%) |
| Employment status | N (%) |
| Paid employee | 2032 (76.7%) |
| Self-employed | 243 (9.2%) |
| On unemployment | 101 (3.8%) |
| Not working - searching for work | 96 (3.6%) |
| On paid leave or furloughed | 41 (1.6%) |
| Retired | 41 (1.6%) |
| Not working - and not looking for a job | 39 (1.4%) |
| On disability or worker's compensation | 35 (1.3%) |
| Other | 22 (0.8%) |
| Job category (multiple choice question) | N (%) |
| Hospital and emergency department workers | 624 (23.5%) |
| Nursing home, long-term care, and home health care workers | 413 (15.6%) |
| Public health workers | 284 (10.7%) |
| Grocery store workers | 283 (10.7%) |
| Teachers and school staff | 251 (9.5%) |
| Food processing workers | 222 (8.4%) |
| Emergency Medical Services workers | 186 (7.0%) |
| Other health care workers | 170 (6.4%) |
| Volunteer (i.e. CERT, MRC, Red Cross, etc.) | 168 (6.3%) |
| Private transportation workers | 156 (5.9%) |
| Sanitation workers | 131 (4.9%) |
| Vaccine manufacturing workers | 121 (4.6%) |
| Postal and shipping workers | 120 (4.5%) |
| Pharmacy workers | 117 (4.4%) |
| Correctional facilities workers | 116 (4.4%) |
| Police or firefighters | 116 (4.4%) |
| Vaccine distribution workers | 95 (3.6%) |
| Other first responders | 93 (3.5%) |
| Public transportation workers | 90 (3.4%) |

### Previous COVID-19 diagnosis, clinical risk, and risk perceptions

As shown in Table 2, twenty four percent of the sample respondents reported having had a prior diagnosis of COVID-19, 83% of whom had no or mild symptoms. Analysis of the clinical risk profile for severe consequences of COVID-19 indicated 26% of respondents reporting one of the seven conditions associated with greater risk, 5% reported two and 2% reported three conditions or more. Fifty-eight percent of respondents were very concerned about getting infected at work, 48% were very concerned about contracting the disease outside the work environment, and 62% were very concerned about the possibility of infecting family members or friends. The factor analysis of these three risk perception questions resulted in one factor with eigenvalue >1, KMO=0.7, alpha=0.8. Based on the factor analysis results, a summative score was created to describe overall risk perception ranging from 0 (low risk) to 6 (high risk), and subsequently three categories of risk perception were created including: low risk perception (up to the 25th percentile), medium risk perception (25th - <75th percentile) and high-risk perception (≥75th percentile). Fifty-five percent of respondents were in the high-risk perception category, 31% in the medium risk category and 14.5% in the low risk.

**Table 2.** Comorbidity, risk perception, experience of discrimination and vaccine hesitancy of the study population

|  |  |
| --- | --- |
| Co-morbidity [diabetes, obesity, rheumatological disease, immunocompromised status, cancer, cardiovascular disease, chronic respiratory disease]. | N (%) |
| No medical condition | 1764 (66.6%) |
| One medical condition | 685 (25.8%) |
| Two or more medical conditions | 201 (7.6%) |
| Have you been diagnosed with COVID-19? | N (%) |
| No | 1961 (74%) |
| I am not sure | 57 (2.2%) |
| Yes, with no symptoms | 266 (10%) |
| Yes, with mild symptoms | 259 (9.8%) |
| Yes, with severe symptoms | 107 (4%) |
| Experience of unfair treatment | N (%) |
| Attributed to any of the following reasons: race, religion, gender and sexual orientation | 1680 (63.4%) |
| Race was the only reason or one of the reasons | 915 (34.5%) |
| Religion was the only reason or one of the reasons | 318 (12%) |
| Gender was the only reason or one of the reasons | 549 (20.7%) |
| Sexual orientation was the only reason or one of the reasons | 361 (13.6%) |
| How concerned are you about any of the following situations ? | N (%) |
| Contracting COVID-19 at work? (*For example: hospital, office, and other work settings that are not your home*) |  |
| Very concerned | 1542 (58.3%) |
| Somewhat concerned | 792 (29.9%) |
| Not concerned | 312 (11.8%) |
| Contracting COVID-19 outside of work? (*For example: at the grocery store, when you are* |  |
| *using transportation, or in other aspects of your daily life)* |  |
| Very concerned | 1266 (47.9%) |
| Somewhat concerned | 1007 (38.1%) |
| Not concerned | 371 (14%) |
| Infecting your family or friends with COVID-19? |  |
| Very concerned | 1653 (62.5%) |
| Somewhat concerned | 664 (25.1%) |
| Not concerned | 326 (12.4%) |
| COVID-19 overall risk perception | N (%) |
| Low risk | 382 (14.5%) |
| Medium risk | 813 (30.1%) |
| High risk | 1439 (54.6%) |
| If you were offered a COVID-19 vaccine within two months from now - at no cost to you- how likely are you to take it? | N (%) |
| Very likely [low hesitancy] | 1059 (40%) |
| Somewhat likely | 523 (19.7%) |
| I would not take it within 2 months but would consider it later on | 188 (7.1%) |
| Not sure | 388 (14.6%) |
| Somewhat unlikely | 153 (5.8%) |
| Very unlikely [high hesitancy] | 339 (12.8%) |

### Past experience with discrimination and vaccine hesitancy

As shown in Table 2, sixty-eight percent of respondents reported having experienced at least once in their lifetime unfair treatment because of their race (34.5%), religion (12%), gender (21%), or sexual orientation (14%). Experience with unfair treatment due to race was reported by all race groups, 62% of Black respondents, 50% of those reporting two or more races, 49% of Hispanic, 45% of Asian, and 25% of white respondents. Experience with unfair treatment due to sexual orientation was reported by 50% of respondents who did not self-identify either as male or female, 23% identified as female and 18% as male. Unfair treatment due to gender was reported by 40% of those self-identifying as neither male or female, 11% identifying as female and 16% as male. In terms of type of experience Black and Hispanic individuals. and those of two or more races reported the most discrimination. *(See Figure 1)*. Forty percent of the sample reported that they would be very likely to take the COVID-19 vaccine, if offered within two months from the time of the survey. In contrast, 13% said they were very unlikely to take it. The remaining forty-seven percent expressed various degrees of hesitancy with 15% responding that they would consider taking the vaccine in the future.

**Figure 1.**
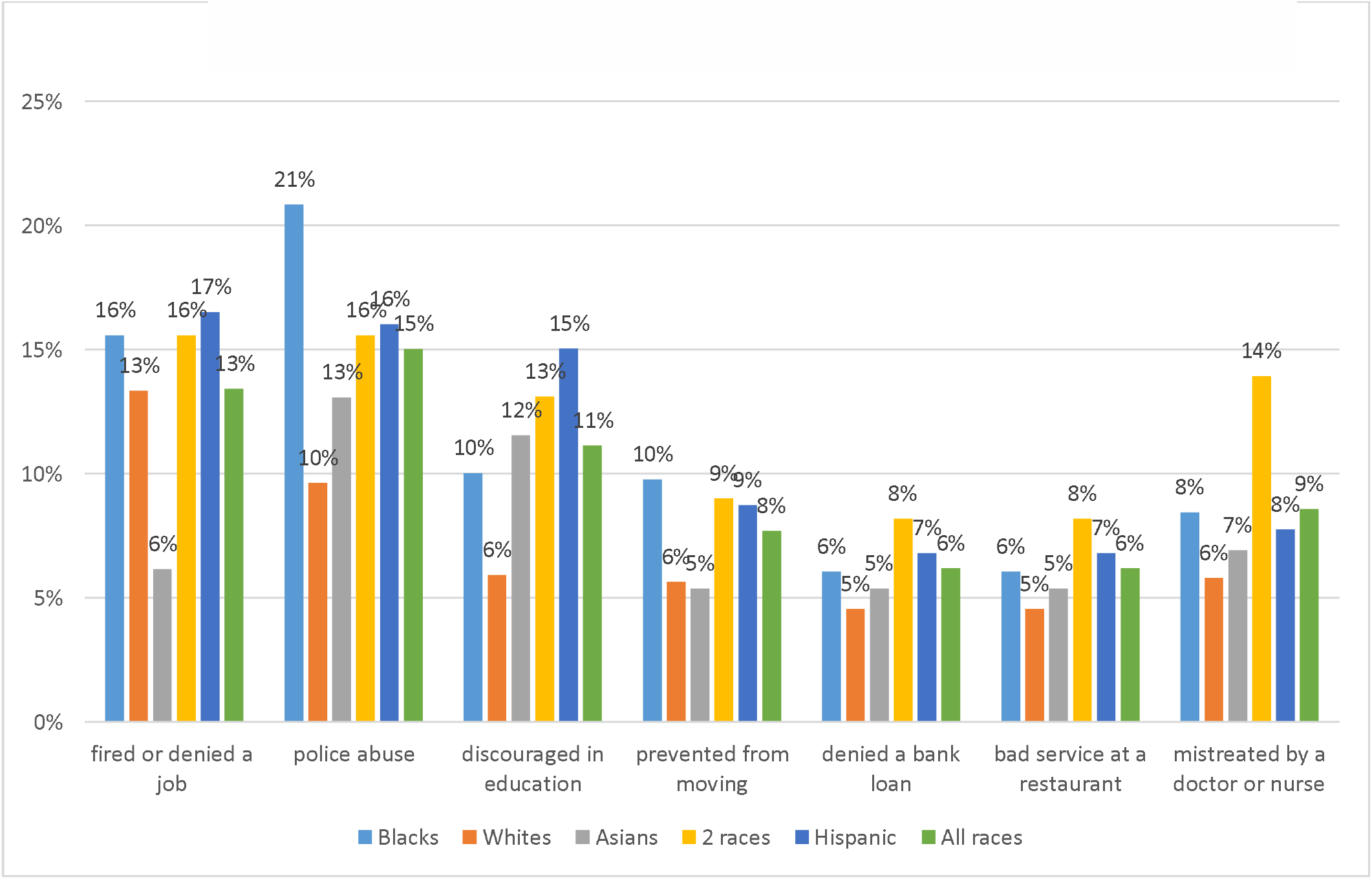
Racial discrimination by type of experience and race.

### Logistic Regression Models

Results of the simple and multivariable regressions are shown in Table 3. In the simple regression models (bivariate analysis) several variables were significantly associated with vaccine hesitancy. Female respondents had 25% decreased odds of reporting a higher level of hesitancy compared to male respondents (OR=0.85, 95% C.I. 0.74-0.98). Respondents with some college education had 34% decreased odds of being at a higher level of hesitancy compared to individuals with less than a high school degree (OR=0.66, 95% C.I. 0.44-0.99). Respondents reporting their race as Black and non-Hispanic had 1.22 times the odds of being at a higher level of hesitancy compared to any other race group (OR=1.22, 95% C.I. 1.01-1.48). Those with a high-risk perception of contracting COVID-19 or of infecting a family member or friend had 1.30 times the odds of being at a higher level of hesitancy compared to those not having such concerns (OR=1.30, 95% C.I. 1.06-1.60). Respondents who had COVID-19 with severe symptoms were more hesitant about taking the vaccine with 1.42 times the odds of being at a higher level of hesitancy compared to those who did not experience the disease at all (OR=1.42, 95% C.I 1.01-1.99). Finally, those who experienced unfair treatment attributed to either their race, religion, gender or sexual orientation had 1.19 the odds of being at a higher level of hesitancy compared to those who did experience discrimination due to the above-mentioned reasons (OR=1.19, 95% C.I. 1.03-1.37).

**Table 3.** Association between independent variables and vaccine hesitancy: simple models and multiple variable models

|  | Simple models |  | Multiple model |  |
| --- | --- | --- | --- | --- |
| Independent variable | OR | 95% C.I. | OR | 95% C.I. |
| Age |  |  |  |  |
| 18-24 | - | - | - | - |
| 25-34 | 1.07 | 0.86-1.34 | - | - |
| 35-44 | 0.99 | 0.80-1.24 | - | - |
| 45-54 | 1.03 | 0.79-1.35 | - | - |
| 55-64 | 0.97 | 0.69-1.36 | - | - |
| 65-74 | 0.75 | 0.43-1.32 | - | - |
| ≥75 | 1.16 | 0.41-3.24 | - | - |
| Gender |  |  |  |  |
| Female *versus* male | 0.85* | 0.74-0.98 | 0.91 | 0.78-1.05 |
| Other than female or male *versus* male | 0.59 | 0.26-1.33 | 0.62 | 0.27-1.42 |
| Employment status |  |  |  |  |
| Paid employee and self-employed *versus* other categories | 1.14 | 0.94-1.39 | - | - |
| Education |  |  |  |  |
| Less than high school | - | - | - | - |
| High school/GED | 0.78 | 0.52-1.15 | 0.78 | 0.52-1.17 |
| Some college | 0.66* | 0.44-0.99 | 0.68 | 0.45-1.01 |
| Bachelor's degree | 0.77 | 0.52-1.13 | 0.78 | 0.52-1.16 |
| Post-graduate degree | 0.97 | 0.66-1.43 | 0.95 | 0.64-1.40 |
| Race |  |  |  |  |
| White non-Hispanic *versus* all other races | 0.94 | 0.81-1.1 | - | - |
| Black non-Hispanic *versus* all other races | 1.22* | 1.00-1.48 | 1.18 | 0.96-1.44 |
| Asian non-Hispanic *versus* all other races | 0.87 | 0.63-1.20 | - | - |
| Hispanic *versus* all other races | 0.92 | 0.71-1.20 | - | - |
| Type of job |  |  |  |  |
| Healthcare sector employee *versus* other job categories | 1.09 | 0.94-1.25 | - | - |
| Medical conditions |  |  |  |  |
| No medical condition | - | - | - | - |
| One medical condition | 1.06 | 0.90-1.24 | - | - |
| Two medical conditions | 1.23 | 0.90-1.69 | - | - |
| Three or more medical conditions | 0.75 | 0.46-1.22 |  |  |
| Risk perception |  |  |  |  |
| Low risk perception | - | - | - | - |
| Medium risk perception | 1.14 | 0.92-1.42 | 1.10 | 0.88-1.38 |
| High risk perception | 1.30* | 1.06-1.60 | 1.18 | 0.95-1.47 |
| COVID-19 diagnosis |  |  |  |  |
| No diagnosis | - | - | - | - |
| Not sure | 1.05 | 0.65-1.68 | 1.01 | 0.62-1.63 |
| Yes – no symptoms | 1.02 | 0.81-1.29 | 0.89 | 0.69-1.13 |
| Yes – mild symptoms | 1.13 | 0.89-1.43 | 1.02 | 0.80-1.29 |
| Yes – severe symptoms | 1.42* | 1.01-1.99 | 1.27 | 0.90-1.79 |
| Experience of unfair treatment |  |  |  |  |
| Attributed to any of the following reasons: race, religion, gender or sexual orientation | 1.19* | 1.03-1.37 | 0.97 | 0.82-1.16 |
| Race was the only reason or one of the reasons | 1.30** | 1.12-1.50 | 1.21* | 1.01-1.45 |
| Religion was the only reason or one of the reasons | 1.21 | 0.98-1.49 | - | - |
| Gender was the only reason or one of the reasons | 0.97 | 0.83-1.14 | - | - |
| Sexual orientation was the only reason or one of the reasons | 0.97 | 0.80-1.19 | - | - |

\* $P < 0.05$
\*\* $P < 0.001$

When the specific reasons for the perceived discrimination were analyzed as independent variables, racial discrimination was the only variable with a significant association with vaccine hesitancy. Those who experienced racial discrimination had 1.3 times the odds of being at a higher level of hesitancy compared to those who had not reported experiencing this type of discrimination (OR=1.30, 95% C.I. 1.12-1.50). *(See Figure 1)*. For the multivariable model, of vaccine hesitancy, the overall LR chi-square test statistics was significant (χ^2^, p<0.01). Brant test p-value resulted 0.68.

In the multivariable models, the only variable associated with vaccine hesitancy was experience of racial discrimination Individuals with past experience had 21% increased odds of being at a higher level of vaccine hesitancy compared to those who did not report such experience. (OR=1.21, 95% C.I. 1.01-1.45). The most frequently reported racial discrimination situation r was abuse from a police officer (15%), followed by having been denied a job or unfairly fired (13%), discouragement in pursuing an education was experienced by 11% of respondents. While all racial groups reported experience with unfair treatment due to their race, Black and Hispanic respondents and those of two or more races reported this experience most frequently.

## Discussion

COVID-19 vaccine hesitancy is developing in a context in which many are fatigued by the mitigation strategies, seeing them as ineffective, and in some cases even punitive. High acceptance of COVID-19 vaccines is critical to ending the pandemic especially among groups for which high transmission rates have been recorded. Based on historical immunization data and current survey results, vaccine hesitancy is higher among Black persons compared to White persons. The low likelihood of getting a COVID-19 vaccine among Black persons is especially concerning because of high rates of transmission in Black communities. Policymakers and public health professionals need to start planning now to make sure the vaccine reaches all Americans — and in particular, that people of color belonging to the priority groups for the vaccination, will not record disproportionally low vaccination rates compared to Whites, and that their concerns will be addressed early on. Public information and warning is one of the preparedness capabilities that public health agencies across the country will need to implement to support the vaccination campaign efforts.^18^ This capability entails the implementation of systems and procedures to mobilize communication activities such as fact gathering, rumor control, message testing, monitoring and publishing content across print, Internet, social, and other media and providing support to spoke persons, such as developing talking points, speeches, and visuals. The results from our study emphasize the need to potentiate monitoring strategies, so to gather information from the public on concerns and reasons for hesitancy towards the COVID-19 vaccine to better target communication efforts to individuals’ informational needs, concerns and experiences. This study has the advantage of focusing on individuals belonging to priority groups to receive the vaccine. This is important as a successful vaccination campaign must demonstrate initial acceptance by the first to be vaccinated. Early adopters of immunization can have a strong influence on the likelihood that others will accept the vaccine and will be compliant with the immunization recommendations. In particular, our sample includes a large fraction of individuals working in the healthcare sector who could be key actors in advocating for the vaccine among the general population. To our knowledge our study is the first to date to include a measure of past experience of discrimination as a predictor of COVID-19 vaccine hesitancy, enriching current knowledge on the relationship vaccine hesitancy and race identified by previous studies. The results show that past-experience of discrimination is a predictor of vaccine hesitancy. This result is important to inform communication and logistical aspects of the COVID-19 vaccination campaign. During the short timeframe of this vaccination campaign, even with the best of intentions, policy makers and public health practitioners will not be able to undo centuries of distrust based on unfair treatment experienced by specific segments of the population in health care, education, finance, and safety. However, they can be sensitive to historical and individuals’ experiences by identifying trusted places and sources of COVID-19 vaccine information and distribution, they can educate clinicians and spokesperson on historical facts, avoiding use of law enforcement to surveil the safety of vaccination sites, engaging individuals from Black communities in vaccination efforts and educating policy makers and vaccine distribution planners on the root causes of mistrust. Enhancing uptake among Black Americans requires much more than disseminating facts about safety - it requires overcoming barriers of mistrust in the system. Policy makers and public officials need to start by acknowledging, appreciating, and discussing mistrust. Labeling those hesitant about the vaccine as conspiracy theorists or individuals unwilling to prevent the spread of the disease, may be counterproductive when hesitancy is rooted in a history of unfair treatment which will not be overturned by denying the existence of fear and doubts. When addressing safety in regards to the COVID-19 vaccine, the content of the message should go beyond the safety of the vaccine per se, and include explicit references and historical comparisons of why this vaccination campaign will not cause another Tuskegee Study. Bidirectional risk communication is of particular importance when one of the goals of the mitigation strategy is to reduce health disparities.

Previous studies have demonstrated the presence of communication inequalities, consisting of inequalities in individual or group specific exposure and reactions to public health communication messages, ^19, 20, 21^ which may lead to further enhance existing disparities across segments of the population in the ability to comply with recommended preventive behaviors. Opinion surveys, at the time of crisis, are a tool to understand people’s concerns so that such concerns can be addressed in communication efforts.^22^ In the case of COVID-19, public officials will need to develop bidirectional communication strategies so that they effectively communicate the correct information concerning the vaccine’s risks and benefits while listening to public opinions and concerns. They also need to be aware that public concerns are not only related to direct health risks due to potential adverse effects from the immunization, but they are also linked to personal beliefs, cultural perspectives and ideology. These concerns are currently further fueled by an emotional dimension driven by the social isolation and daily life restrictions and difficulties brought on by the pandemic. Public health agencies need to enhance their public information capabilities to address multiple dimensions of the vaccine communication strategy in order to be successful and meet growing needs of information, reassurance and address mistrust to the extent possible.

### Study Limitations

Because we used a cross sectional study design, the timing of the survey must be considered in interpreting and generalizing the results. The survey was fielded in December 2020 when vaccines were announced but not yet available to the public. Due to the evolving epidemiology of the disease, and developing public communication and vaccine distribution efforts the predictors of vaccine hesitancy are likely to change overtime, in particular in regards to the impact of some independent variables for which, in our study, we did not find a statistical significant association with vaccine hesitancy such as risk perception of contracting COVID-19. Our sample is not a representative sample of the US population as such study results are not generalizable outside the study population. While our sample included a distribution of racial-ethnic groups that allowed us to analyze predictors of vaccine hesitancy based on race it did not include a sufficient number of individuals over 65 which based on previous studies are more likely than others to accept the COVID-19 vaccine due the increased risk of severity in the elderly.

## Conclusions

Results from this survey of a convenience sample of the US population show that past experience with discrimination is a predictor of vaccine hesitancy. This result is important to inform communication and logistical aspects during the COVID-19 vaccination campaign which need to be sensitive to individuals’ past experience with systemic unfair treatment by different types of institutions including law enforcement, education and healthcare.

## Supporting information

Appendix - Survey Instrument

STROBE checklist

## Data Availability

Data will be available upon request

