## Appendix - Survey Instrument for "Predictors of COVID-19 Vaccine Hesitancy: Socio-demographics, Co-Morbidity and Past Experience of Racial Discrimination"

HARVARD T.H. CHAN SCHOOL OF PUBLIC HEALTH
Emergency Preparedness Research, Evaluation, and Practice Program

### Vaccine Hesitancy Survey

December 2020

For reproduction contact

* 1. **Are you a worker in any of the following categories?**

❑ Hospital and emergency department workers

❑ Nursing home, long-term care, and home health care workers

❑ Public health workers

❑ Emergency Medical Services workers

❑ Correctional facilities workers

❑ Sanitation workers

❑ Vaccine manufacturing workers

❑ Vaccine distribution workers

❑ Other health care workers

❑ Pharmacy workers

❑ Teachers and school staff (including childcare and K-12)

❑ Food processing workers

❑ Grocery store workers

❑ Postal and shipping workers

❑ Public transportation workers

❑ Private transportation workers

- ❑ Police or firefighters
- ❑ Other first responders
- ❑ Volunteer (i.e. CERT, MRC, Red Cross, etc.)
- ❑ Other (please specify)

*Screening question

#### Tell us about you ....

* 2. **Do you work in the healthcare sector?**

- ○ Yes
- ○ No

Continue to tell us about you ....

* 3. **Which title <u>best</u> represents you?**

- ○ Physician (MD or DO only) or Physician Assistant (PA)
- ○ Nurse (RN), Nurse Practitioner (NP), Certified Nurse Midwife (CNM), or Other Nurse Professional (e.g. Licensed Practical Nurse (LPN), Certified Nursing Assistants/Aides (CNA), etc.)
- ○ Dentist or Other Dental Professional
- ○ Pharmacist
- ○ Social Worker/Mental Health Professional
- ○ Other (please specify)

* 4. **What is your age category?**

- ○ 18-24
- ○ 25-34
- ○ 35-44
- ○ 45-54
- ○ 55-64
- ○ 65-74
- ○ 75+

* 5. **What is your gender?**

- ○ Male
- ○ Female
- ○ Prefer to specify (please specify)

* 6. **In what state or U.S. territory do you live? [if you do not live in the USA select the option "other country" at the end of the list]**

7. **What is your zip code? (use 00000 for outside the U.S.)**

* 8. **What language(s) do you usually speak at home? [check all that apply]**

- ❑ English
- ❑ Spanish
- ❑ French or French Creole
- ❑ Vietnamese
- ❑ Filipino
- ❑ Portuguese or Portuguese Creole
- ❑ Chinese
- ❑ Other (please specify)

* 9. **What race/ethnicity do you consider yourself?**

- ○ White, Non-Hispanic
- ○ Black, Non-Hispanic
- ○ Asian, Non-Hispanic
- ○ 2+ races
- ○ Hispanic
- ○ Prefer not to say
- ○ Other (please specify)

* 10. **What is the highest level of schooling you have completed?**

- ○ Less than high school
- ○ High school/GED
- ○ Some college
- ○ Bachelor's degree
- ○ Post-graduate degree (i.e. Master, PhD, MD, etc)
- ○ Other (please specify)

* 11. **Select the employment status that <u>best describes</u> your current situation [select one option only]:**

- ○ I am working - paid employee
- ○ I am working - self-employed
- ○ I am not working - on unemployment
- ○ I am not working - on paid leave or furloughed
- ○ I am not working - searching for work
- ○ I am retired
- ○ I am not working - on disability or worker's comp
- ○ I am not working - and not looking for a job
- ○ Other (please specify)

* 12. **During the past 12 weeks did you experience difficulties in affording food or medical care?**

- ○ Yes
- ○ No

* 13. **Do you have any of the following conditions ? [check all that apply]**

- ❑ Cancer
- ❑ Severe allergies
- ❑ Seizures
- ❑ Immunocompromised state due to therapy or disease
- ❑ Autoimmune disease
- ❑ Obesity
- ❑ Diabetes (type 1 or 2)
- ❑ Cardiovascular disease
- ❑ Pulmonary disease
- ❑ Rheumatological condition
- ❑ Pregnancy

❑ I do not have any medical condition
❑ Other (please specify)

* 14. **How concerned are you about any of the following situations?:**

| | Very concerned | Somewhat concerned | Not concerned |
|---|---|---|---|
| **Contracting COVID-19 at work?** (For example: hospital, office, and other work settings that are not your home) | ○ | ○ | ○ |
| **Contracting COVID-19 outside of work?** (For example: at the grocery store, when you are using transportation, or in other aspects of your daily life) | ○ | ○ | ○ |
| **Infecting your family or friends with COVID-19?** | ○ | ○ | ○ |

#### Your experience ....

* 15. **Have you been diagnosed with COVID-19?**

* 16. **Did any of your close family members or friends experience any of the following? [Check all that apply]:**

❑ Tested positive for COVID-19 and had no symptoms or or mild symptoms
❑ Tested positive for COVID-19 and had severe symptoms
❑ Died of COVID-19
❑ Lost their job or had a salary reduction due to COVID-19
❑ None of my close family or friends experience any of the above situations

* 17. **We are interested in the way other people have treated you. Can you tell us if any of the following situations ever happened to you at any time in your life?: [Check all that apply]**

❑ You were unfairly fired or denied a job
❑ You were unfairly stopped, searched, questioned, physically threatened or abused by the police
❑ You were unfairly discouraged by a teacher or advisor from continuing your education
❑ You were unfairly prevented from moving into a neighborhood because the landlord or a realtor refused to sell or rent you a house or apartment

- ❑ You were unfairly denied a bank loan
- ❑ You received a service at a restaurant that was worse than what other people received
- ❑ You were unfairly treated by a doctor or nurse
- ❑ None of the above situations have ever happened to me

18. **If any of the above situations happened to you do you think it was due to any of the following reasons?**

- ❑ Your race
- ❑ Your sexual orientation
- ❑ Your gender
- ❑ Your religion
- ❑ Other reason (please specify)

* 19. **What do you think about the number of cases of COVID-19 reported in your state?**

- ○ The number of cases being reported is <u>much lower</u> than the actual number of cases
- ○ The number of cases being reported is <u>much greater</u> than the actual number of cases
- ○ The number of cases being reported is <u>somewhat accurate</u>
- ○ I don't know

* 20. **Have you received the Flu vaccine this year? [check only one option]**

- ○ Yes
- ○ No, but I will get it
- ○ No, and do not plan to get it (explain why)

21. **Were you ever recommended a vaccine by a healthcare provider that you did not take?**

- ○ Yes
- ○ No
- ○ I do not remember

22. **If you did not take the vaccine that was recommended to you, what was/were the reason(s)? [check all reasons that applied to that situation]**

- ❑ I did not think it was necessary
- ❑ I did not have enough information about the vaccine
- ❑ I did not think the vaccine was effective
- ❑ The vaccine was too expensive
- ❑ It was not logistically convenient
- ❑ I did not think the vaccine was safe
- ❑ I was concerned about the side effects

- ❏ I had a prior bad experience with vaccinations
- ❏ I was afraid of needles
- ❏ For religious reasons
- ❏ Other (please specify)

* 23. **Tell us how well the following statements describe your reactions and thoughts:**

| | Very concerned | Somewhat concerned | Not concerned |
|---|---|---|---|
| I feel anxious when I see the number of COVID-19 cases climbing | ◯ | ◯ | ◯ |
| I find the prospect of a vaccine exciting | ◯ | ◯ | ◯ |
| I feel depressed about the uncertainty of how this pandemic will evolve | ◯ | ◯ | ◯ |
| I get upset when I hear contradictory information about COVID-19 | ◯ | ◯ | ◯ |
| I feel stressed when I am unable to plan my life due to COVID-19 | ◯ | ◯ | ◯ |
| I think that taking chances is part of life and so is taking the vaccine | ◯ | ◯ | ◯ |

#### Information about the vaccine ...

* 24. **Have you heard about a COVID-19 vaccine**

- ○ Yes, and it was mostly positive
- ○ Yes, and it was mostly negative
- ○ Yes, and it was neither positive or negative
- ○ No

...continue on information

* 25. **Did you get information about the COVID-19 vaccine from social media ? [check all that apply]**

- ❏ No, I did not
- ❏ I am not sure
- ❏ Yes - from Facebook
- ❏ Yes - from YouTube
- ❏ Yes - from Instagram
- ❏ Yes - from TikTok
- ❏ Yes - from Twitter
- ❏ Other social media (please specify which one)

#### Social media use ...

26. **Did the information you got from <u>social media</u> change your level of confidence in the COVID-19 vaccine?**

- ○ Increased my confidence in the vaccine
- ○ Decreased my confidence in the vaccine
- ○ Did not change my confidence
- ○ I am not sure
- ○ Did not change my confidence but influenced my opinion in other ways - please specify

27. **Did you ever share information on <u>social media</u> about the COVID-19 vaccine?**

- ○ Yes
- ○ No
- ○ I do not remember

.... trust in information

* 28. **Where did you get the most information about the COVID-19 vaccine? Select up to 3 SOURCES:**

- ❑ Local television news (on TV or on the web)
- ❑ English language national or cable network news (on TV or on the web)
- ❑ Non-English language television station (on TV or on the web)
- ❑ National newspaper (i.e. New York Times, Wall Street Journal, USA Today on paper or on the web)
- ❑ My town or other local newspaper (on paper or on the web)
- ❑ Non-English language newspaper (on paper or on the web)
- ❑ English language radio
- ❑ Non-English language radio
- ❑ News portal site such as Yahoo! or MSN
- ❑ Website of a government agency
- ❑ Social media
- ❑ Word of mouth
- ❑ Through my employer
- ❑ Other (please specify)

* 29. **How much do you <u>trust the information</u> you got so far about the COVID-19 vaccine?**

- ○ Not at all
- ○ Very little
- ○ Somewhat
- ○ A lot

* 30. **Who would you trust the most to give you information about the COVID-19 vaccine in the near future? Select your <u>TOP 3 choices</u>:**

- ❏ Federal officials
- ❏ Your state officials
- ❏ Your town leaders (mayor or board of selectmen)
- ❏ Public health experts
- ❏ Your employer
- ❏ Your co-workers
- ❏ Your doctor
- ❏ Your local pharmacy
- ❏ Your family and friends
- ❏ Your community health center
- ❏ A celebrity (for example: a sports figure, actor, or musician)
- ❏ Local leaders in my community not in government positions (for example: local organizations, religious leaders)
- ❏ Other (please specify)

#### Your opinions ...

* 31. **If you were offered a COVID-19 vaccine within two months from now - at no cost to you- how likely are you to take it?**

- ○ Very likely
- ○ Somewhat likely
- ○ I am not sure
- ○ Somewhat unlikely
- ○ Very unlikely
- ○ I would not take it within 2 months but would consider it later on

* 32. **Select the top 3 locations you would trust the most to get the COVID-19 vaccine:**

- ❑ Doctor’s office
- ❑ Local pharmacy
- ❑ Urgent care center
- ❑ Hospital
- ❑ Community health center
- ❑ Local health department
- ❑ School
- ❑ Church
- ❑ Military facility
- ❑ Community center
- ❑ Local park or community outdoor space
- ❑ Any location
- ❑ None of the above
- ❑ Other (please specify)

confidence

* 33. **What would be important for you to know to make you more likely to take the COVID-19 vaccine? <u>Select up to 3 options</u>**

- ❏ The fast production of the vaccine did not compromise its safety
- ❏ Those approving the vaccines are following strict rules
- ❏ My risk of getting sick with COVID-19 is bigger than the risk of side effects from the vaccine
- ❏ The vaccine cannot cause any immediate or long term injury
- ❏ It is impossible to get COVID-19 or any other disease from the vaccine itself or its components
- ❏ The vaccine works in protecting me from COVID-19
- ❏ The vaccine works in stopping the transmission of COVID-19 from one person to another
- ❏ The FDA, CDC, and WHO recommend the vaccine and agree it is safe
- ❏ Other (please specify)

* 34. **What else ...... would be important for you to know to make you more likely to take the COVID-19 vaccine? <u>Select up to 3 options</u>**

- ❏ Once vaccinated I will be able to live my life with no restrictions
- ❏ Those with concerns about the vaccine have opportunities to share their opinions with the public
- ❏ Pharmaceutical companies will not make large profits from the vaccine
- ❏ Everybody will have equal access to the vaccine regardless of income, race, or insurance status
- ❏ I will be free to choose if I get the vaccine or not with no consequences
- ❏ There is no other reason why we have so many people sick (i.e. 5G technology or other factors we do not know about)
- ❏ Other (please specify)

35. **If you have other opinions about the vaccine you would like to share please write them here;**

Thank you !

**Thank you for taking this survey!**
